# Time-series ECG Imputation Using a Pattern-Based Masking Framework

**DOI:** 10.64898/2026.01.14.26344164

**Authors:** Sukardi Suba, Alexander Novak, Xiaojuan Xia, Salah S. Al-Zaiti, Michele M. Pelter

## Abstract

The utilization of continuous ECG monitoring has become an integral part of modern hospital-based care. However, missing data presents significant challenges in deploying real-time ECG-based predictive systems. Research on the implementation of imputation techniques on time-series ECG is limited. Furthermore, the performance of imputation techniques is typically benchmarked using random masking, which may not reflect the real-world missingness patterns encountered in clinical practice. This study aimed to evaluate and benchmark a range of imputation methods, from conventional statistical approaches to state-of-the-art deep learning models, using continuous ECG time-series data under different missingness conditions, including both random (conventional) and observed pattern-based (realistic) missingness.

Time-domain features were extracted from continuous 12-lead Holter recordings (ranging from 2.5 to 4 hours per patient) from a pilot cohort of 40 patients. Missingness was introduced using random and pattern-based masking. We compared seven imputation methods: global mean, linear interpolation, *K*-*N*earest *N*eighbors (KNN), *M*ultiple *I*mputation by *C*hained *E*quations (MICE), softImpute, xgboo*S*t *MI*ssing va*L*ues in tim*E S*eries (SMILES), and *S*elf-*A*ttention-based *I*mputation for *T*ime *S*eries (SAITS). Performance was evaluated using mean absolute error (MAE) across masking approaches, missingness levels, and missingness patterns.

Overall, the MAEs for all seven imputations are higher under pattern-based masking than random masking. SAITS achieved the best performance across both masking approaches (MAEs of 0.277 and 0.146; standard deviations of absolute error of 0.398 and 0.252 for observed-pattern and random masking, respectively). Simpler methods such as SoftImpute and KNN showed comparable performance across both masking approaches, and particularly under certain missingness levels.

Artificially masking by random may underestimate the accuracy of time-series imputation in real-world scenarios. Our findings underscore the importance of context-based imputation strategies (i.e., masking approach and imputation method) and balancing model complexity with practical considerations (e.g., resources, costs, and level of missingness) for real-time deployment.

**Author Summary:** Missing data is a common issue in healthcare research, especially in continuous recordings such as electrocardiograms (ECGs), which are used to monitor heart activity over time. When certain measurements are missing, researchers and clinicians often rely on *imputation, which is simply a technique* to“fill in the blanks” However, how well imputations work can depend on *why* and *how* the data went missing in the first place.

In this study, we compared several imputation methods, ranging from basic averaging to advanced machine learning techniques, using real patient ECG data. To better reflect the types of missingness observed in clinical settings, we applied a new approach to simulate more realistic data gaps and patterns. We found that some of the more complex models performed better overall, although simpler methods were surprisingly resilient in certain situations.

Our findings underscore the importance of aligning the imputation strategy with the specific data challenges. Our work may help guide future use of ECG data in real-time prediction models and improve how missing information is handled in healthcare research and practice.

## 1. Introduction

Continuous electrocardiographic (ECG) monitoring has become increasingly integral to modern hospital-based clinical practice and research,^1^ especially as healthcare systems move toward proactive, predictive, and artificial intelligence (AI)-enabled models of care.^2^ Real-time ECG waveforms can serve as a rich, temporally resolved data source for detecting and forecasting a wide range of clinical conditions, including arrhythmias,^3–6^ cardiorespiratory decompensation,^1,7,8^ and myocardial ischemia.^9,10^ These capabilities are particularly relevant in high-acuity settings such as intensive care units, step-down/progressive care units, and telemetry units, where continuous ECG and other physiologic (e.g., vital signs) surveillance is standard of care.

However, a major challenge in deploying real-time predictive systems based on ECG signals is the high rate of missing data. In the real-world clinical setting, continuous ECG recordings are frequently disrupted due to both patient– and equipment-related factors. Common causes include electrode displacement due to patient movement or mobility therapy, poor skin-electrode contact caused by sweat, body hair, or compromised skin integrity, and hardware issues such as lead detachment or connectivity loss during transfers for diagnostic testing or procedures.^11–13^ These disruptions can lead to short or extended intervals of missingness, ranging from milliseconds to several minutes, or even >15 minutes, as reported in our previous work,^14^ and often occur in non-random, clinically meaningful contexts.

The impact of missing data on time-series analyses, particularly when applying machine learning models for clinical data forecasting, is well established.^15,16^ While various imputation techniques have been developed and widely applied to time-series data derived from electronic health records (EHRs) data,^17–19^ most of this work has focused on structured tabular data (e.g., lab values, vital signs).^16^ Far less attention has been paid to continuous ECG data, which pose unique challenges due to their high sampling rate, periodicity, and vulnerability to transient or sustained artifacts.^20–22^

Recent advances in time-series imputation, including deep learning methods such as BRITS (Bidirectional Recurrent Imputation for Time Series)^23^ and SAITS (Self-Attention-based Imputation for Time Series),^24^ offer promising approaches for reconstructing missing values in complex, multivariate time-series. However, these models have seldom been benchmarked specifically using ECG data. Physiologic signals, particularly ECG, require imputations that not only minimize error metrics but also preserve clinically relevant waveform features, such as the QRS complex, ST-segments, and T-waves, which are essential for downstream diagnostic tasks.

Furthermore, many published imputation studies rely on datasets with artificial or uniformly random missingness patterns, which may not accurately reflect the clinically driven, realistic missingness encountered in ECG monitoring.^16,21,22,25^ Additionally, to the best of our knowledge, the effects of missing value types (*random* versus *not random*) have not been extensively studied in the context of time series data imputation.^21^ Understanding how different imputation techniques perform under such conditions is crucial for developing robust AI systems that can function reliably in real-time clinical environments. This is especially important for ensuring that models used in real-time monitoring systems remain resilient to data loss while maintaining predictive performance.

Therefore, to address this gap, this study aimed to evaluate and benchmark a range of imputation methods, from conventional statistical approaches to state-of-the-art deep learning models, using continuous ECG time-series data under different missingness conditions, including both random (conventional) and observed pattern-based (realistic) missingness.

## 2. Methods

### 2.1. ECG Data Source

Twelve-lead Holter recordings (H12+, Mortara Instrument, Milwaukee, WI) from the COMPARE Study were used. Briefly, the COMPARE Study was a prospective observational study designed to examine transient myocardial ischemia (TMI) in hospitalized patients with suspected non-ST-segment elevation acute coronary syndrome (NSTE-ACS). The design, methods and inclusion/exclusion criteria have been described in detail in previously published papers.^11,26^ Research nurses obtained signed informed consent prior to application of the Holter recorder, which used the Mason-Likar limb lead configuration. To ensure high-quality ECG recordings a standardized protocol was used (i.e., thorough skin preparation, chest hair clipping as necessary, and high-quality radiolucent skin electrodes so as to not interfere with X-rays).^11^ The research nurses made frequent rounds throughout the day to maintain accurate electrode placement and re-application of electrodes when indicated (i.e., electrode(s) that had fallen off, post echocardiogram, X-ray, or invasive coronary angiography). The 12-lead ECG Holter data was collected in the context of a research protocol (e.g., black box) and was not used for clinical decision making; hence, hospital-based ECG monitoring (bedside or telemetry) was maintained per eachhospital’s protocol. The mean time from hospital admission to application of the 12-lead ECG Holter recorder was six hours (± 5 hours), and recordings were maintained until hospital discharge or the patient went to open heart surgery.

### 2.2. ECG Pre-Processing and Feature Extraction

Twelve-lead ECG Holter signals were collected using a high-fidelity flashcard capable of recording 1,000 samples/second/channel. For the analysis reported here, we included available ECG recordings from 40 hospitalized patients who were admitted with non-ST-segment elevation chest pain.

We focused on extracting time-domain ECG features from a 2.5 – 4-hour continuous segment of leads II, III, aVF, V3, and V4. These leads were specifically chosen to balance computational cost and feasibility with signal quality, as well as to align with the clinical focus of the study. Leads II, III, and aVF represent the inferior region of the heart, while V3 and V4 are anterior precordial leads. These territories are frequently evaluated for myocardial ischemia or infarction, which is central to our pilot investigation. In over half of the cases, waveform segments required manual adjudication due to signal artifacts, pacing, or chronic arrhythmias such as atrial fibrillation. These issues often necessitated the exclusion or substitution of cases, making analysis of the entire 12 leads infeasible. Limiting the analysis to five leads allowed for consistent, high-quality feature extraction within a manageable time frame of our pilot study.

The selected 2.5 – 4-hour window also informed by the real-world practice, mimicking the approximate time needed to rule-in/-out ACS in the emergency department (ED). For patients in the ACS group, the analyzed segment was drawn from the period leading up to the pre-identified ischemic events, which were annotated for their true events in the primary study (COMPARE Study)^26^. For the non-ACS group, we used the initial hours of Holter data acquired upon ED arrival. This window allowed for consistent clinical framing across groups while accommodating the practical constraints of data quality and manual signal validation. The ECG features extracted included RR intervals (*RR*), QRS duration (*QRSd*), QRS amplitude (*QRSamp*), QRS area (*QRSarea*), QT interval (*QT*), heart rate-corrected QT (*QTc*), ST-segments at the J-point plus 60– and 80-milliseconds (*ST-J60 and ST-J80, respectively)*, T-wave amplitude (*Tamp*), T-wave length (*TLen*), and T-wave morphology (*Tanno*: *1-Positive, 2-Low Amplitude, 3-Biphasic, 4-Negative*).

Time-domain ECG features were extracted using a 10-beat median beat signal averaging approach. For each patient, continuous ECG signals were segmented into rolling windows of ten consecutive, valid QRS complexes. Candidate windows were selected based on RR interval (within 10% variation of RR median). This helped exclude ectopic beats (e.g., premature atrial or ventricular complexes) but did not fully eliminate noisy or non-analyzable beats. As a result, some windows lacked sufficient signal quality for feature extraction and were recorded as missing. Within each window, a median beat waveform was constructed by aligning individual sinus beats and selecting the pointwise median voltage across time. The features were then calculated from this representative median beat. The analysis proceeded in a rolling, non-overlapping manner with each 10-beat window advancing sequentially through the recording. In some instances, analyzable beats might be <10 due to noise (poor signal quality) and non-sinus beats. We recorded the number of analyzable beats for each window in each row.

### 2.3. Imputation Pipeline and Methodology

#### 2.3.1. Missing Data Simulation for Imputation Evaluation

Missing ECG data from continuous recording are typically not missing at random. We observed temporal dependencies with extended periods of missingness for certain features, as well as cross-feature dependencies, especially for features derived from shared waveform segments. For example, the QT interval cannot be measured if either the QRS onset or the T-wave offset is unclear, directly linking its availability to the quality of those segments.

Missingness in this study refers to the absence of extracted feature values for a given lead at a given time point, as determined by the signal averaging and quality control process described above. Specifically, a feature value was marked as missing if a valid 10-beat median waveform could not be constructed due to insufficient analyzable QRS complexes, typically resulting from signal noise, pacing artifacts, or rhythm irregularities. Although missing values could result from raw lead dropout or complete disconnection of electrodes, we intentionally excluded such cases for this pilot study given its limited scale. We also did not attempt to reconstruct any lead using other leads (e.g., viaEinthoven’s law or vectorcardiographic inference). Each lead-feature pair was evaluated independently based on its own waveform availability.

To understand and evaluate how different imputation methods respond to realistic time-series ECG-specific missing data patterns, we artificially introduced missing values into a low-missingness ECG dataset to serve as a controlled ground truth. Known values were masked and subsequently imputed to allow us to compare imputation results to the original data and compute error metrics. We conducted two trials of masking to simulate missing data: (1) Masking by Observed Patterns, and (2) Masking by Random. The former provided a more realistic evaluation of imputation performance on ECG data, while the latter provided a commonly used^16,21,22,25^ but more naive baseline for the performance of the examined imputation algorithms.

##### a) Masking by Observed Patterns

To simulate a more realistic, non-random missingness pattern, we designed and implemented a *transfer-based masking* approach, where we applied the missingness pattern from a patient with higher natural missing values to a patient with low missingness, effectively transferring the observed gaps to a more complete dataset. To conduct this trial, we first identified a“base patien” with the lowest percentage of naturally missing values. Thepatient’s data frame (defined as a two-dimensional tabular structure with timepoints as rows and features as columns) contained 51 numerical columns (ECG features) and 1,678 time steps (rows), totaling 85,578 data points, of which only 243 (0.28%) were naturally missing. After normalization (zero mean, unit variance per feature), this patient served as the ground truth reference for performance evaluation.

We then created a set of“mask source patients” defined as those without any fully missing columns (features) from whom missingness patterns were taken to create the masking applied to the base patient. Therefore, for each base–source patient pair, we applied the exact missingness pattern from the mask source patient to the basepatient’s data frame, effectively transferring the mask. Specifically, the missing values were introduced to the basepatient’s data frame in the same locations as the missing values in the mask sourcepatient’s data frame. That is, if a particular value was missing at time (row) ***t*** for a feature (column) ***f***, in the mask source patient, the corresponding value at ***t,f*** location in the base patient was masked. The original values in these locations were saved to compute performance metrics after imputation at the later steps. This approach allowed us to simulate realistic missingness while retaining the ground truth value underneath for error analysis.

If the base and mask source patient data frames differed in length (i.e., number of rows), both were truncated to the shorter length. We excluded any base–mask source patient combinations that resulted in fully missing columns after the masking process. This step yielded a final set of 39 data frames with different mask variations.

##### b) Random Masking Trial

In this trial, the artificially missing values in the mask were introduced uniformly *at random*, a commonly used technique for imputation validation.^16,21,22,25^ The random masking trial had a similar process to that of the masking by observed patterns above. However, the actual pattern of missingness (i.e., which feature columns and time step rows were missing) from the mask source patient data frame was not preserved. Instead, for each selected patient pair, the missing values were introduced into the basepatient’s data frame at randomly selected locations, but at a rate equal to the overall proportion (percentage) of missing data observed in the corresponding mask source patient. To ensure consistency and fair comparisons with the masking by observed pattern trial above, we also excluded the same base patient–mask source patient pairs that resulted in fully missing columns in the masking by observed pattern trial. This trial also resulted in a set of 39 masked data frames. An illustration of both masking trials is presented in **Fig 1**.

**Fig 1.**
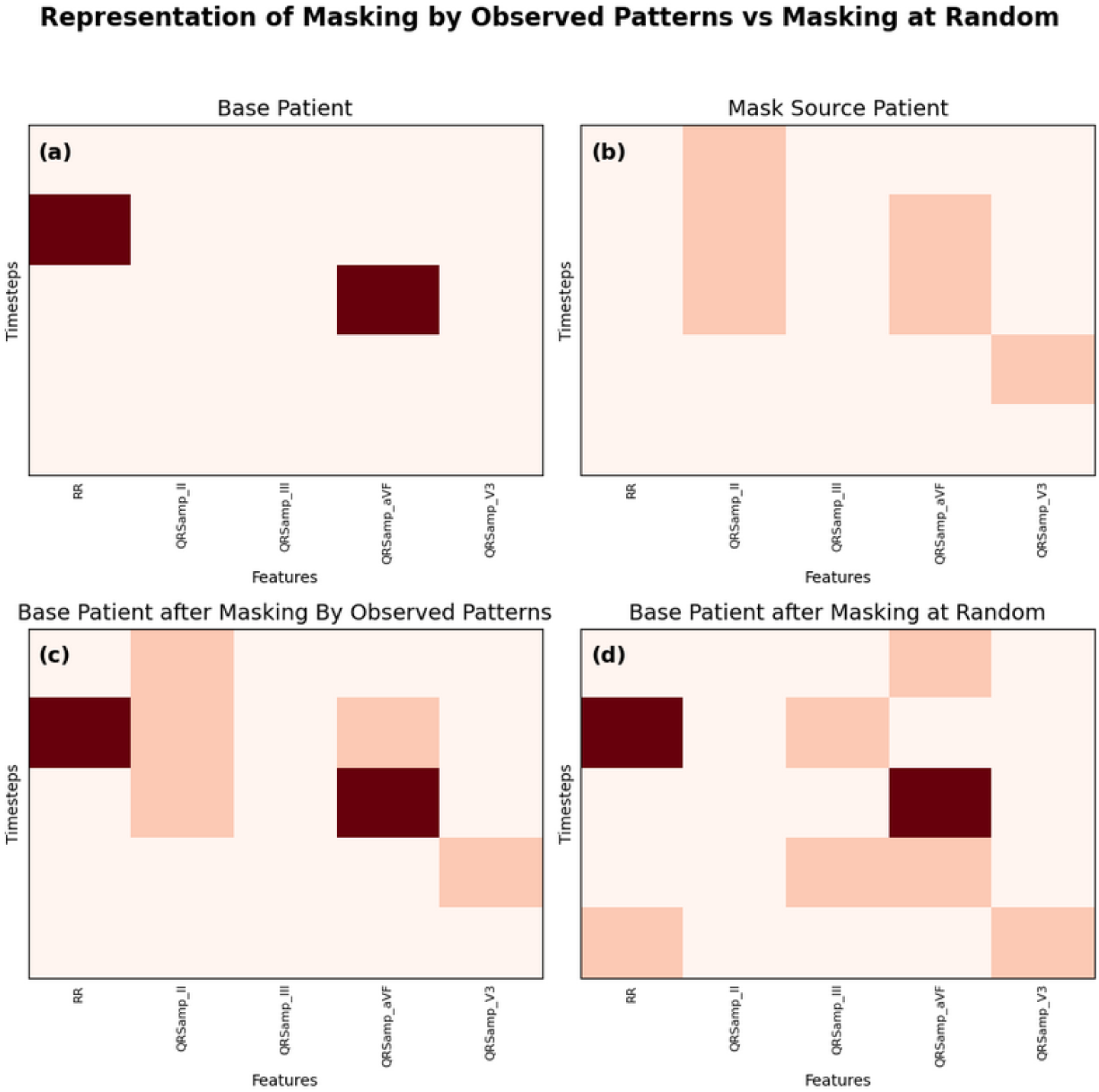
Visual representation of the two different masking trials on example data frames. The off-white/pale peach color (all charts) represents observed (non-missing) values. **Top Charts: (a)** Base Patient with minimal naturally missing data (0.28% of the entire data points). The dark red squares represent the naturally missing data. **(b)** Mask Source Patient. The dark peach squares indicate the location of the missing values for that patient, which then was used as a pattern for artificial masking on the Base Patient (Masking by Observed Patterns Trial). **Bottom Charts: (c)** Base Patient + Transferred Mask from Source. The colors represent the combination of the artificially introduced missingness pattern (dark peach squares) from (b), which then being used to assess imputation performance, and the naturally missing values (dark red squares) of (a), which cannot be used in imputation performance calculations. **(d)** Base Patient + Random Mask (Same % Missing). The chart shows the artificially masking values (dark peach squares) based on the same percentage of missing values as in Mask Source Patient. The dark red also indicates the naturally missing values of the Base Patient.

#### 2.3.2. Imputation Methodologies

We then applied several methods to estimate the values of the artificially introduced missing values in our dataset to evaluate the effectiveness of various imputation strategies. These methods represent a range of complexities, from simple statistical techniques traditionally used in time-series analysis to modern, state-of-the-art models which have only been developed in recent years.

When discussing and evaluating imputation strategies, it is important to distinguish between two types of patterns in the data. *Temporal* patterns refer to how one feature of a dataset varies (or changes) over time within a subject. *Spatial* patterns refer to the relationship between multiple features at a single time point. Some imputation methods will rely solely on spatial patterns, treating each timestep independently without considering the time series nature of the data. Others may only examine one feature at a time during imputation based on its past and future values, taking advantage solely of temporal patterns. Some of the most advanced models may leverage both temporal and spatial dependencies simultaneously. Below we describe several methods, from simple statistical to more advanced methods.

##### a) Baseline Statistical Imputation

The two simplest imputation algorithms that we examined as reference points for model performance were linear interpolation and the global mean method. In global mean imputation, missing values for a given feature are filled using the mean of all non-missing values for that feature. This method is simple and fast, and serves as a common baseline method for imputing missing values.^27^ Another simple method that may perform better by having a greater focus on the time series structure of the data is linear interpolation.^27^ In linear interpolation, missing values for each feature are estimated by interpolating linearly between the nearest previous and subsequent non-missing values in the time series. Both methods are univariate in nature and apply imputations by only considering information from one feature at a time (feature-by-feature). They do not utilize cross-feature correlations or spatial structure. Their performance in our analysis served as a baseline when comparing more complex and advanced imputation methods.

##### b) Spatial-Only Imputation Methods

We implemented two multivariate, spatially-aware imputation strategies using the *scikit-learn* library. First was the K-Nearest Neighbors (KNN) method,^28^ which we implemented through*sklearn’s KNNImputer*.^29^ In this method, for some set value of ***k***, the KNN algorithm imputes a missing value of a certain feature at a particular timestep by determining the ***k most similar timesteps*** in the dataset where the specified feature is observed. The similarity of the two timesteps (the *missing value* and the potential *k-nearest* timesteps) is determined using the Euclidean distance across other available features, not considering any with missing values. The imputed value is then obtained as a weighted average (based on similarity) of the observed values from the *k* nearest neighbors. Thus, this method utilizes the additional context from all features in the dataset to perform imputations. In our imputation tests, we used ***k=10***, selected by varying the ***k*** and evaluating the resulting performance across features with different missingness rates (**Suppl. Fig 1**). Our goal was to select a value of ***k*** that would provide a balance of good performance across all levels of missingness.

The second method was Multivariate Imputation via Chained Equations (MICE),^30,31^ which we implemented through *IterativeImputer*.^32^ The MICE imputation is an iterative multivariate regression-based technique. Initially, all missing values were filled using the global mean method as discussed previously. Then, for each feature in turn, missing values were imputed (or predicted) by fitting a BayesianRidge regression model trained on the remaining features, including previously imputed ones. This process was repeated for a certain number of iterations, and the predictions from the last round were used as the final imputed values. Using a similar process as with tuning the KNN***“”*** parameter, we set the number of iterations for the MICE imputation method to be 320 (**Suppl. Fig 2**). The two methods above (KNN and MICE) exploited cross-feature correlations but did not account for the time-series structure of the data.

##### c) Temporal AND Spatial Imputation

The remaining two imputation methods we examined considered the time series nature of the dataset, where the temporal proximity of observations provided additional context that might be useful when predicting missing values. We first evaluated a modified version of the SMILES (xgbooSt MIssing vaLues In timE Series) framework,^33^ which uses XGBoost models trained on a temporally localized input window to impute missing values. In our adaptation of this imputation technique, we first prefilled all missing values in the dataset using the softImpute^34^ matrix completion algorithm, implemented via the *fancyimpute*^35^ package. The parameter controlling the maximum number of iterations for the softImpute algorithm was also tuned in a similar fashion to the KNN and MICE parameters (**Suppl. Fig 3).**

The softImpute algorithm approximates a provided data matrix with a low-rank representation by iteratively applying Singular Value Decompositions (SVDs). In each iteration, the singular values are shrunk by a regularization parameter lambda (***λ***) and then thresholded at zero to prevent negative values. This iterative shrinkage allows the method to recover latent structure in the data while handling missingness. We used the default method of filling the missing values before performing the SVDs, which initially sets all missing values to zero. The regularization parameter ***λ*** was also left at its default setting, which would set it to the maximum singular value in the initial matrix divided by 50. We set the maximum number of iterations to 1000 and replaced the originally missing values in our dataset with the corresponding values from the completed low-rank matrix produced by softImpute. This prefilled dataset served as the input for the XGBoost-based imputation in the SMILES framework.

We then trained extreme gradient boosting (XGBoost)^36^ models, as implemented in the *DMLC xgboost* library,^36^ to predict the originally missing values for each feature. A separate model was trained per feature, with each model learning to impute any originally missing values of its target feature by leveraging temporally localized input data, including the prefilled data, from all features.

In our implementation, we constructed a training dataset `(X, y)` where each output `y` represented an originally non-missing value of the target feature. The corresponding input `X` comprised all values, across all features, within a fixed temporal window (three timesteps) centered on the same timestep. Specifically, for a value `y` located at timestep *t*, the input `X` consisted of values from all features between timesteps *t – 3* and *t + 3*. Any originally missing values within this window were substituted with their prefilled softImpute estimates from the prior step above. Once trained, the XGBoost model was used to predict the originally missing values of the target feature, using the same sliding-window input format at each missing timestep.

During the implementation process, we found that the model performance was highly sensitive to several hyperparameters. These parameters are heavily context dependent, and their optimal values must be specified: maximum tree depth (`max_depth`), the shrinkage rate for feature weights (`learning_rate`), and the proportion of samples used for training each tree (`subsample`). Specifying them improperly could lead to large declines in model performance. To combat this in our SMILES implementation, we conducted a grid search for each feature-specific XGBoost model, varying `max_depth` ∈ {2, 4, 6}, `learning_rate` ∈ {0.05, 0.1}, and `subsample` ∈ {0.8, 1}. Finally, the masked values were evenly split into a testing and a validation set, with the final performance metrics computed only on the held-out testing set.

For completeness, we also included the softImpute algorithm predictions alone as a standalone imputation method (i.e., without the XGBoost post-processing step). We used the same implementation and settings as in the SMILES prefilling step described previously.

The last imputation strategy we implemented was Self-Attention-based Imputation for Time Series (SAITS) method,^24^ a deep learning model designed to capture both temporal and spatial dependencies among the data during imputation using Diagonally Masked Self-Attention (DMSA) blocks. While the SAITS method does allow the deep learning model to be trained on multiple samples of data at once, we trained the SAITS model separately for each patient to remain consistent with other methods and isolate patient-level variation.

We used the implementation provided in the *PyPOTS* library^37^ and performed a grid search to determine the optimal values for the following hyperparameters:

- number of layers in the DMSA blocks: `n_layers` ∈ `{1,2,3}`
- the input dimension of the multi-head DMSA layers: `d_model` ∈ `{64,128,256,512}`
- the number of heads in the DMSA mechanism: `n_heads` ∈ `{2,4,8}`, and
- the dimension of the layers in the feed-forward networks: `d_ffn` either set to the same value as `d_model` or half the value of `d_model`.

In all cases, a dropout of 0.1 was used to prevent overfitting, which we later confirmed to be sufficient for this task as there was no evidence of any overfitting in graphs of the validation performance of SAITS by training epoch.

The best-performing configuration was `n_layers = 3`, `d_model = 512`, `n_heads = 8`, and `d_ffn = 512`. However, we set `n_layers = 1` for the remainder of our experiment to reduce training time and minimize cost for computational needs since it had minimal impact on performance. To further guard against overfitting, we also used a validation set to trigger early stopping during the training process. The best model was then selected based on validation performance, using a validation set constructed in a similar fashion as described previously for the SMILES model. Again, the final imputation performance metrics were computed only on the testing set.

We summarize all seven imputation techniques, their utilization of temporal and spatial patterns, underlying imputation strategies, and publication year in **Table 1**.

**Table 1.** Comparison of imputation methods by modeling approach and temporal context.

| Imputation Method | Utilizes Spatial Patterns | Utilizes Temporal Patterns | Technique Used | Year Published |
| --- | --- | --- | --- | --- |
| Global mean |  |  | Basic Computation | Unclear (*commonly used statistical method*)<sup>38</sup> |
| Linear Interpolation |  | X | Basic Computation | Unclear (*widely available in software libraries*)<sup>39</sup> |
| KNN | X |  | Distance Based | 1951<sup>28</sup> |
| MICE | X |  | Regression Based | 2000<sup>30</sup> |
| SoftImpute | X | X | Matrix Factorization | 2010<sup>40</sup> |
| SMILES | X | X | Tree-based | 2020<sup>33</sup> |
| SAITS | X | X | Neural Network | 2023<sup>24</sup> |

Note: *KNN*, K-Nearest Neighbor; *MICE*, Multivariate Imputation by Chained Equations; *SAITS*, Self-Attention-based Imputation for Time Series; *SMILES*, xgbooSt MIssing vaLues in timE Series.

### 2.4. Comparison of the Imputation Technique

The primary metric used to evaluate the performance of each imputation method was mean absolute error (MAE):

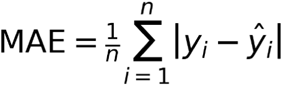

This formula describes how to determine the MAE of the set of *n* predicted values 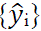 based on the corresponding set of true values {*y_i_*}.

We calculated MAE-based metrics for all experiments described below to evaluate performance across imputation methods and masking strategies. As a baseline performance metric for each imputation method, we computed the MAE for the imputations of all the artificially introduced missing values in our dataset, as well as the standard deviation of the absolute errors. Additionally, we computed feature-specific MAE for a representative subset of the features to provide a granular insight into method performance across physiologically related variables. We selected all ST-segment subset features as this representative subset because our focus domain is examining acute myocardial ischemia. We further generated bar charts with error bars representing 95% confidence interval (CI) for the MAE within each feature based on the imputation methods and masking strategies.

We also examined the MAEs to understand how the imputation methods performed based on two aspects of data missingness patterns. The first aspect was the proportion of missing values by column. We hypothesized that imputation methods leveraging temporal or spatial patterns might struggle when data is extensively missing along the corresponding dimension. To test this, we binned each imputed value according to the proportion of missing values (both natural and artificially introduced) in their feature column and calculated the MAE within each bin. We repeated the same process using the proportion of missing values by row to examine the impact of timestep-level missingness.

The second aspect of missingness patterns we examined was the“gap lengt” of an imputed value to nearest non-missing observation. We defined gap length as the minimum number of consecutive missing values preceding or following a certain missing value; essentially, its distance to the nearest non-missing value. Similarly to the comparison analyses discussed previously, we binned each imputed value according to its gap length and then computed the MAE within each bin.

To increase reproducibility of our work, all codes of our framework presented here are available on GitHub at https://github.com/suksuba/ecg-imputation.

## 3. Results

Across the 40 patients included in the analysis, eachpatient’s data frame contained 54 feature columns. The dataset comprised a total of 80,698 timesteps (rows), resulting in 4,357,692 individual data points. On average, each patient contributed 1,876.7 rows, with a range from 922 to 3,068. Across all patients, the average number of QRS complexes used for signal averaging within 10-beat window was 9.4, with individual patient means ranging from 4.15 to 9.95. The minimum number of analyzable beats used for constructing a median beat in any given window was two, and the maximum was ten.

The performance metrics of the seven imputation methods for both missingness trials (masking by observed patterns and random masking) are presented in **Table 2**. These metrics were computed based on all the imputed values from the 39 masked data frames pooled together. The standard deviation (SD) columns indicate the SD of the absolute error for an individual imputed cell.

**Table 2.** Summary of the mean absolute error (MAE) and standard deviation (SD) of absolute error metrics for all seven imputation methods for both masking trials.

| Imputation Methods | Random Masking |  | Masking by Observed Patterns |  |
| --- | --- | --- | --- | --- |
|  | MAE | SD of Absolute Error | MAE | SD of Absolute Error |
| Global Mean | 0.559 | 0.795 | 0.673 | 0.772 |
| Linear Interpolation | 0.249 | 0.387 | 0.446 | 0.583 |
| KNN | 0.184 | 0.272 | 0.326 | 0.442 |
| MICE | 0.127 | 0.389 | 0.350 | 1.544 |
| SoftImpute | 0.149 | 0.246 | 0.331 | 0.440 |
| SMILES | 0.124 | 0.227 | 0.304 | 0.429 |
| SAITS | 0.146 | 0.252 | 0.277 | 0.398 |

Across all methods, the MAE was higher when missing values were artificially introduced using observed patterns compared to random masking. The magnitude of the absolute error increase varied by method. The global mean method showed the smallest change, with an MAE increase of 20.3% (from 0.559 to 0.673). In contrast, MICE showed the largest change, with a 241.1% increase (from 0.127 to 0.350). In most cases, the standard deviation of the absolute error also increased under observed-pattern masking, indicating greater variability in imputation performance. The only exception was, again, the global mean method, which showed a slight decrease (from 0.795 to 0.772). These results indicate that the simpler methods were more resilient to the missingness pattern applied, and thus, were less impacted by the masking approach. Structured missingness posed a greater challenge for more complex approaches.

We also examined the MAEs achieved for each imputation method on specific features. **Fig 2** shows the performance of each method on a subset of ST-segment features, which include ST-segment measured at J-point + 60 msec (STJ60) and at J + 80 msec (STJ80) for each of the five ECG leads (II, III, aVF, V3, and V4). In both masking trials, the performance of imputation methods varied greatly by feature, except for the global mean method which achieved a similar MAE for all features. In general, while both masking trials seemed to broadly agree on which features were comparatively more or less difficult to impute, the MAEs reported for a specific feature did differ. For example, both masking trials indicated that SAITS performed better when imputing the *STJ60_V3* feature than the *STJ60_II* feature, although the exact MAE values for both were somewhat different.

**Fig 2.**
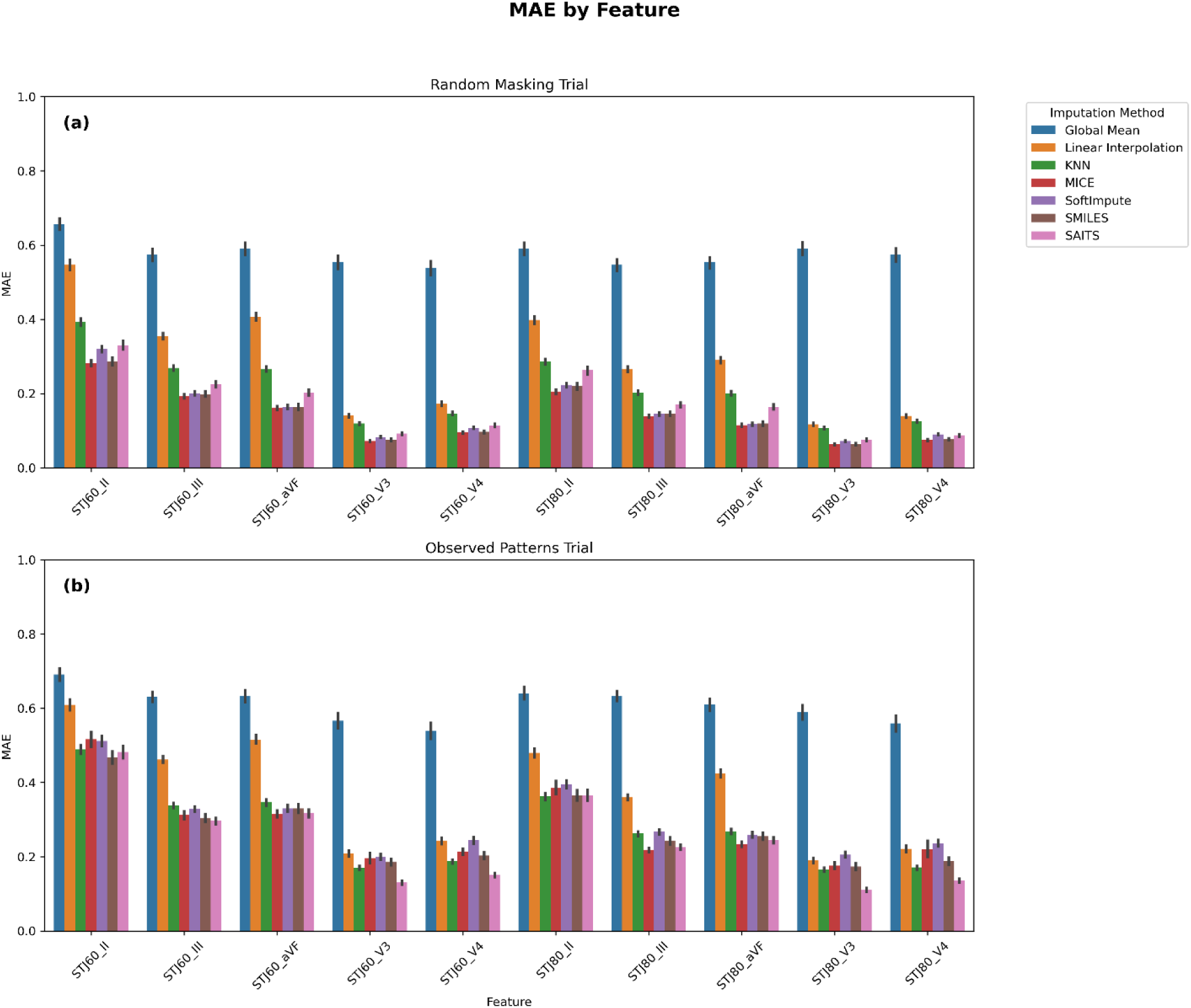
Overall imputation method performance (MAE) by feature for each masking trial. **(a)** masking by random, and **(b)** masking by observed patterns.

The MAEs achieved from each imputation method according to the percentage of missing data in the targetcell’s feature *column* (i.e., missingness by column) are shown in **Fig 3**. Note that we did not observe any columns formed with more than 50% missing values under the random masking trial. Thus, those bins are omitted from the chart. The error bars (vertical lines on top of each bar) represent 95% confidence intervals for the true mean absolute error of imputed values in each bin. In the random masking trial, the imputation methods did not see much change as the proportion of missing data by column increased. At each level of missingness, the global mean performed the worst followed by the linear interpolation method. The rest of the methods performed similarly well except the KNN method, which had slightly worse performance across all levels of missingness.

**Fig 3.**
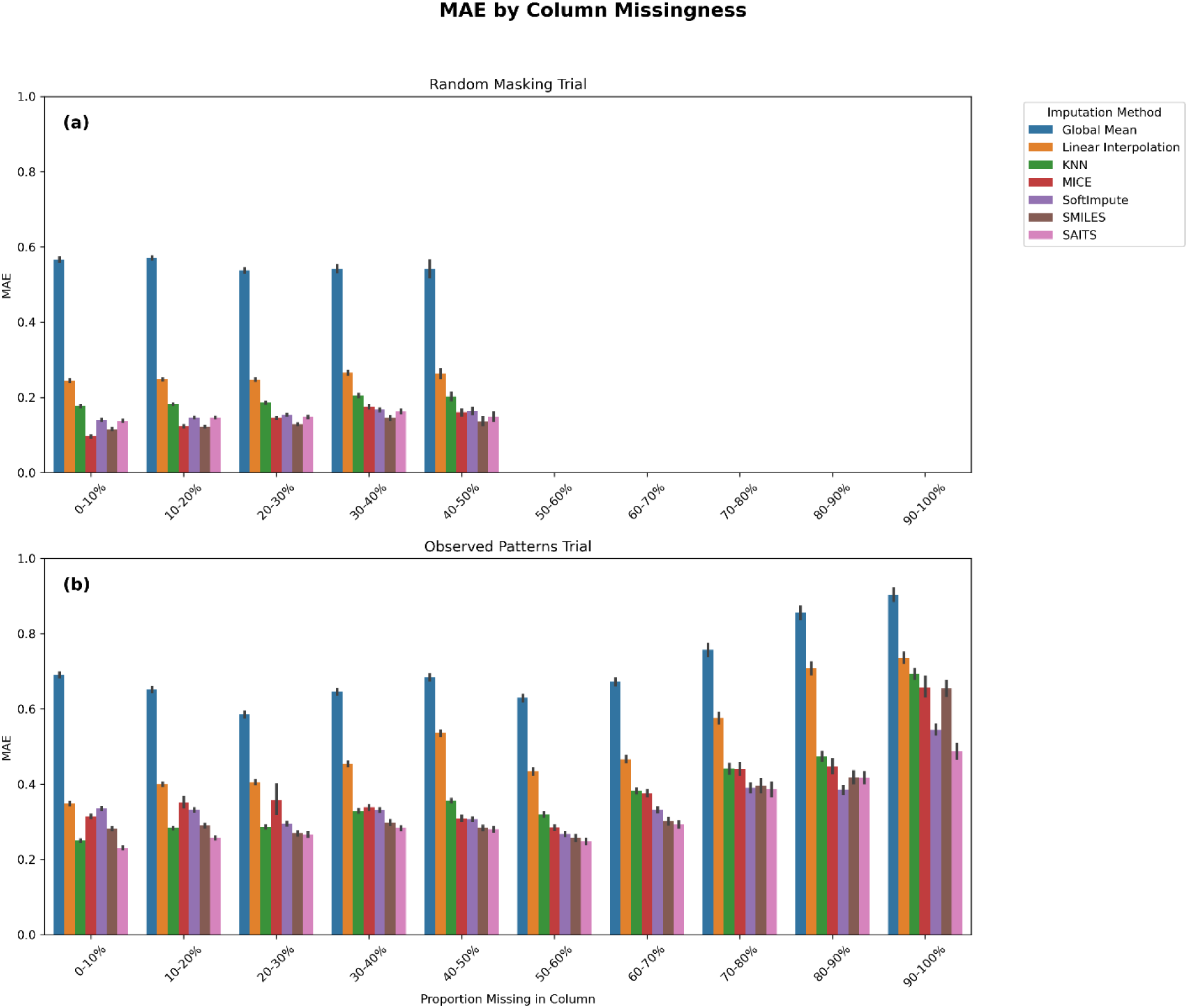
Imputation method performance by proportion of missing data in column for each masking trial. (a) masking by random, and (b) masking by observed patterns.

In the masking by observed patterns trial, the magnitude of the MAEs achieved by all imputation methods (except the global mean) was greater than the MAE achieved in the random masking trial for all levels of missingness. In addition, we saw more changes in the performance of each method at the differing levels of missingness. In general, we noted allmethods’ performance somewhat worsened as the proportion of missing data in a column increased. When only ≤10% of the data was missing in a feature column, SAITS performed best, followed by KNN. However, as the proportion of missingness increased to the 50-60% range,KNN’s performance worsened further than SAITS, SoftImpute, and SMILES, all of which achieved a similar MAE. While it was difficult to draw conclusions due to wider error bars, SAITS notably appeared to perform best when the proportion of missing data was high (≥60%). MICE performed comparatively poorly when the proportions of missing data were low, generally achieving similar MAE as simple linear interpolation. However, MICE was more resilient to higher proportions of missing data, generally outperforming linear interpolation when the proportion of missing data was ≥40%. Similar to linear interpolation, the global mean method also suffered some declines in performance for greater levels of missingness.

The MAEs achieved by each imputation method based on the percentage of missing data by row of the target cells are shown **Fig 4**. Note that in the random masking trial, there were no rows formed that had more than 80% of the data missing, and in the masking by observed patterns trial, there were no rows formed that had more than 90% of the data missing. Therefore, these bins were left blank on the charts. The error bars indicate 95% confidence intervals for the true mean absolute error of a cell imputed by a certain method and located in a row with the appropriate proportion of missing data. These error bars are extremely large in the 60-70% and 70-80% bins of the random masking due to low sample size.

**Fig 4.**
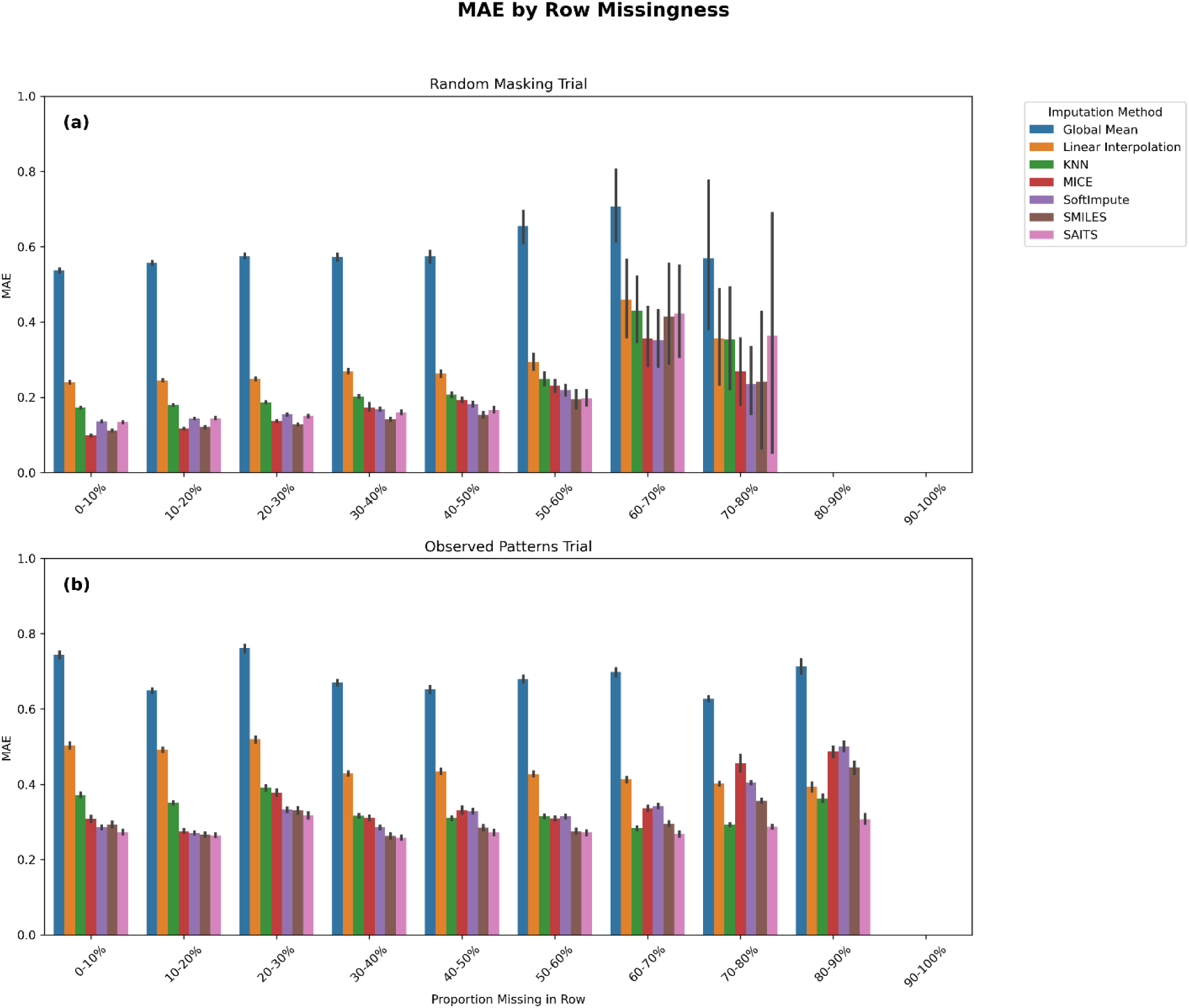
Imputation method performance by proportion of missing data in row for each masking trial. (a) masking by random, and (b) masking by observed patterns.

In the masking at random, most imputation methods exhibited some increase in MAE as the proportion of missing data by row increased. However, it was difficult to determine the magnitude of this effect due to the wide confidence intervals in the 60-70% and 70-80% bins. If we excluded these two bins, due to the potential unreliability of their results, some specific patterns related to the imputation method did emerge. The MICE method was heavily impacted by the amount of data missing in a row, as it initially performed the best among all the methods but was outperformed by the more resilient SAITS and SMILES in the 30-60% missing data range. The linear interpolation and global mean methods exhibited little change across the different bins, which was not surprising, as they did not account for spatial patterns. The KNN, SoftImpute, and SMILES methods exhibited remarkably similar changes in performance across these different levels of missingness, with SMILES consistently outperforming SoftImpute, which in turn outperformed KNN.

However, in the masking by observed patterns trial, most imputation methods did not show a significant change in MAE as the proportion of missing data by row increased. Generally, if a consistent decline in performance occurred, it was only in the bins with the highest amounts of missing data. Even the MICE imputation method, which showed very clear signs of declining performance as missing data grew more prevalent in the masking at random trial, did not show a similar pattern in this trial. Again, we noted that the magnitude of the MAEs was greater across all levels of missingness by row in the masking by observed patterns trial (compared to random masking) for most imputation methods.

One factor that might help explain the differences in performance between trials, at least for certain imputation methods, was the presence of long gaps of missing data in the masking by observed patterns trial. **Fig 5** illustrates how some of these gaps appear in a selectedpatient’s data frame. Each column represents a different feature, and each row represents a particular timestep, with the red dashes indicating a missing value in that location within the data frame. We would expect these extended periods of consecutive missing values in one feature not to appear when masking at random, as supported by **Fig 6**. In the masking at random trial, most of the gaps created were less than four timesteps long, and thus, we pooled observations with gap length of 4 or more into a single bin (**Fig 6A**). However, we observed that the consecutive missingness in the masking by observed patterns trial could result in very long gaps of missing values, with some extending over 200 timesteps (**Fig 6B**).

**Fig 5.**
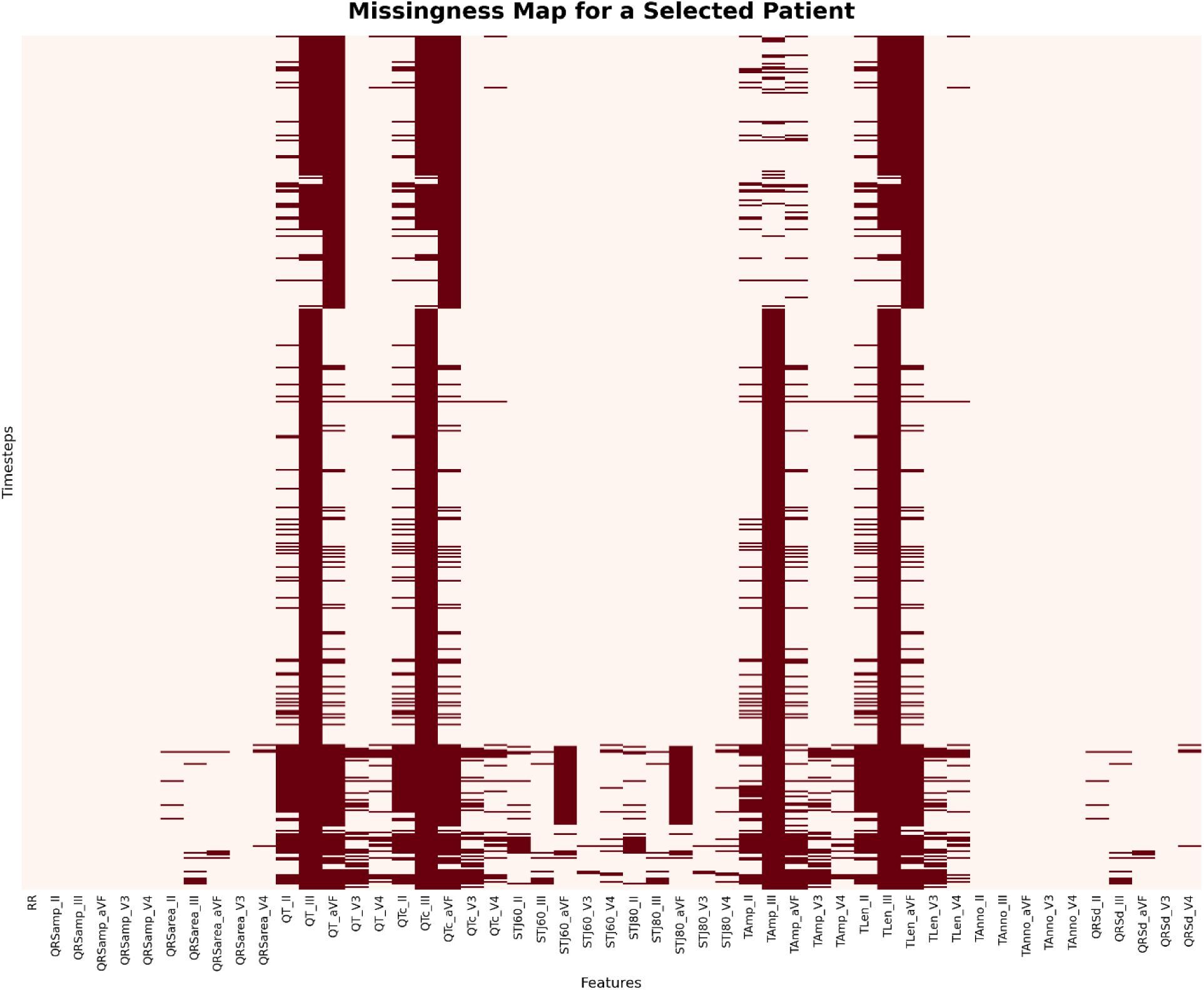
An example plot showing the locations of missing values (dark red color) in a patient’s data frame. The X-axis shows the ECG features (i.e., column variables in the data frame), and the Y-axis indicates the time step (i.e., row in the data frame). Note the continuously long, vertical dark red “lines” for QT_III, QTc_III, Tamp_III, and TLen_III, which indicate many consecutive missing values. This plot also shows a great example of spatially correlated features. The QT interval of the ECG is measured from the onset of the QRS complex to the offset of the T-wave. Unclear boundaries of the T-wave (TLen) lead to an unmeasurable QT interval, which dictates how corrected QT (QTc) is calculated. T-wave amplitude (Tamp) may also affect the T-wave’s boundaries.

**Fig 6.**
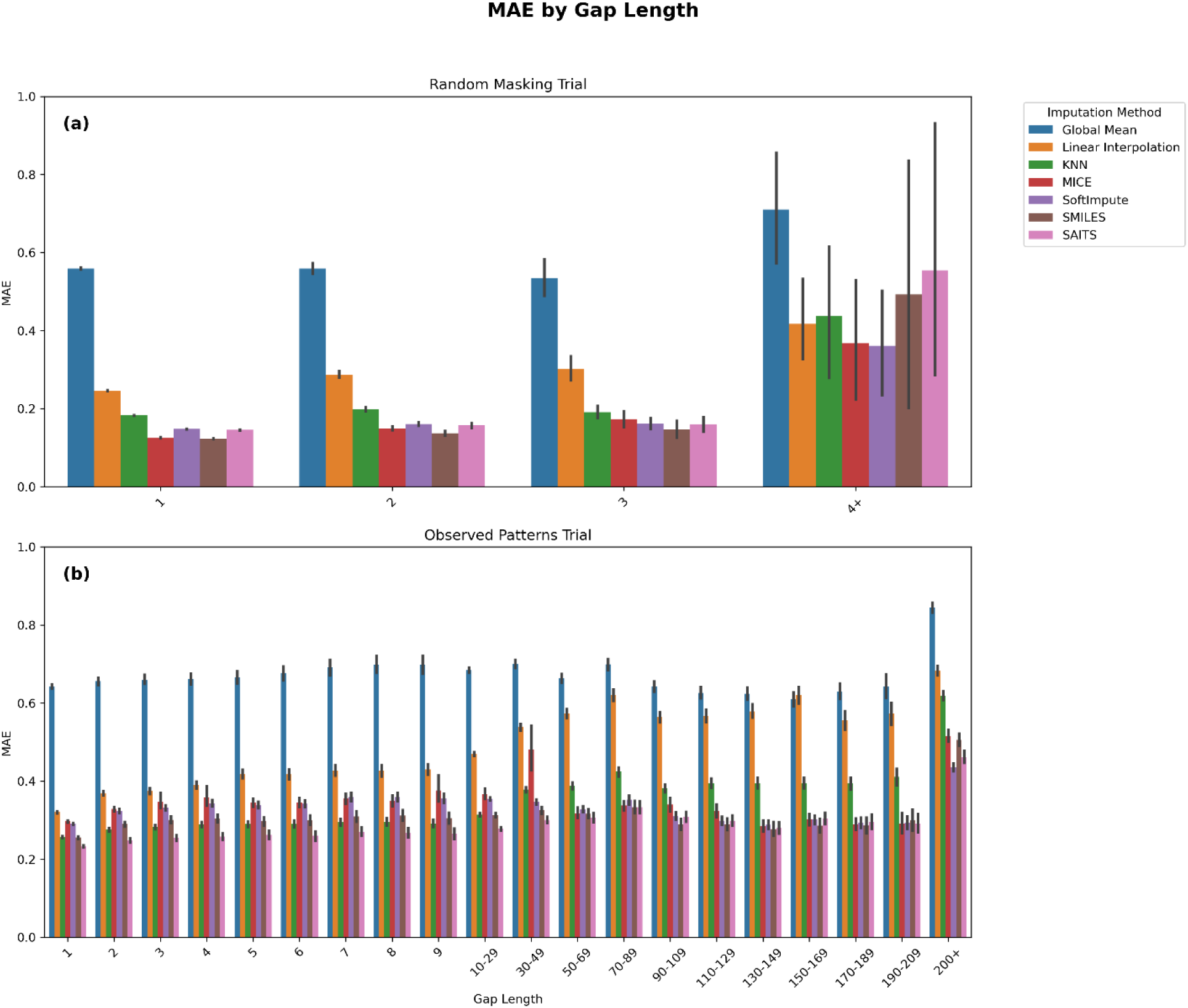
Comparison of imputation methods performance using the mean absolute error (MAE) by gap length for. **(a)** random masking and **(b)** masking by observed patterns. Note that random masking generated a few missingness gaps beyond 4. Vertical lines on top of each bar indicate error bars representing a 95% confidence interval.

Despite the extensive missingness gaps introduced in the masking by observed patterns trial, many of the imputation methods we tested did not seem to be strongly impacted by these gap patterns. Linear interpolation, which relied solely on nearby temporal patterns to impute missing values, showed an increase in MAE as the gap length increased. Most other imputation methods did not show a similar increase in MAEs for the longer gap lengths. We would expect imputation methods such as MICE and the global mean method not to be impacted by gap length, as they do not rely on temporal information for their imputations. However, even methods such as SAITS and SMILES did not appear to be affected by gap length until the gap exceeded 200 consecutive missing values.

## 4. Discussion

We implemented conventional statistical-based (linear interpolation and global mean), machine learning-based (KNN, MICE, SoftImpute, and SMILES), and deep learning-based (SAITS) imputation methods on continuous, time-series ECG data from 40 patients. We further evaluated and benchmarked the performance of the seven imputation methods by applying observed pattern-based masking approach against random masking (conventional approach).

Overall, the two masking trials that we tested showed markedly different benchmarking outcomes for the seven imputation algorithms. We argue that masking by observed patterns more closely approximates real-world missingness in ECG measurement datasets, as it reflects a more realistic missingness distribution. Therefore, comparing performance across the two masking trials provides insights into how commonly used random masking might misrepresent true imputation performance in time-series ECG data.

Across nearly all imputation methods, the MAE was substantially higher when evaluated using observed patterns of missingness. In some cases, the MAEs doubled or even tripled the estimates relative to that when using random masking. All methods more sophisticated than global mean imputation showed at least a 77.3% increase in MAE under observed-pattern masking. This suggests that evaluations relying solely on random masking may significantly underestimate real-world error. This underestimation would be difficult to correct, given that the magnitude of the MAE inflation varied substantially between methods. In addition to overall error, the variability of the absolute error for most imputation methods also increased under the observed patterns masking trial, based on the standard deviation of the absolute error. This suggests that random masking would also underestimate the variability of the error associated with the imputed values on ECG data.

Random masking also limits the conclusions that can be drawn when examining imputation performance relative to different missingness patterns (i.e., missingness by row or column), as we showed in **Figs 3** and **4**. For instance, the random masking trial did not produce any feature columns with >50% missing values (**Fig 3**), precluding performance evaluation of the imputation techniques under high column-wise sparsity. This is especially important to consider when analyzing ECG-derived features, which often result in high levels of column-wise missingness. Furthermore, even within low-to-moderate missingness ranges, the two masking trials showed different outcomes. As shown in **Fig 3**, the MAEs under random masking remained relatively stable (or changed very minimally) as the proportion of column-wise missingness increased. In contrast, the observed-pattern trial showed more dynamic changes, with error rates fluctuating across different levels of missingness and with the relative performance of imputation methods shifting accordingly. An opposite trend was seen in the case of row-wise missingness, as shown in **Fig 4**. We hypothesize that this is due to the fact that the most correlated features tend to go missing at the same time, and therefore additional missingness in a row is unlikely to impact the imputation error very much. However, under random masking, interpretations became more challenging when levels of missingness were high due to small sample sizes.

We can further speculate on why certain imputation techniques are more sensitive to the underlying pattern of missingness. Note that the global mean method was the least affected, which is not surprising, given its simplicity makes it relatively immune to structural distribution of missing data. The only plausible mechanism for degraded performance in this method would be the increased presence of columns with extremely high missingness in the observed-pattern masking trial, which could reduce the accuracy of the estimated values. However, this effect appears to be minimal, as the MAE increased by only 20.3%, which suggests that the global mean remained relatively stable even when the column-wise sparsity was high.

Masking based on observed patterns, as opposed to random masking, can challenge imputation techniques that rely on either temporal or spatial information. Observed-patterns masking may often introduce long consecutive gaps of missing values, which can impair methods that depend on nearby values for accurate prediction. Additionally, if correlated features tend to be missing simultaneously at the same time step, spatial dependencies may also be disrupted.

Longer gaps appear to mostly strongly affect methods that rely solely on temporal continuity. This can be clearly seen in linear interpolation method, which utilizes only temporal values (rather than spatial ones) to fill in missing data. As shown in **Fig 6**, its MAE increases with increasing missing data gaps length, although it plateaus beyond a gap length of 90. Despite this, linear interpolation was one of the methods least impacted by the masking style, with a 79.6% increase in MAE when masking by observed patterns (see **Table 2**), likely because it always relies on immediate neighbors rather than a wider temporal or spatial context.

In contrast, more advanced temporally-aware methods like SAITS, SoftImpute, and SMILES showed relatively stable performance across gaps length until the largest “200”) bin (**Fig 6**). Small increases in MAE were observed when moving from a gap length of 1 (where at least one *immediately* adjacent timestep, either directly before or after the missing value, contains a known value for the same feature) to longer gaps. This is likely because these methods tend to rely primarily on temporally close values (nearby timesteps) when performing imputation. For example, the XGBoost component of the SMILES framework is limited to only using data within three timesteps (±3 timesteps window) of the missing data for its imputation.

While it is possible that observed masking patterns interfere with temporal context in other subtle ways, it seems plausible that the bulk of the MAE increase for these imputation methods is attributed to disruption of spatial information. That is, the concurrent absence of correlated features at the same timestep likely undermines the spatial dependencies that these models also leverage.

The MICE imputation method, which relies solely on spatial information to fill missing values, was particularly vulnerable to structured missingness under the observed-patterns masking. Its MAE more than tripled, increasing by 276.8% compared to when using random masking. This substantial performance decline suggests thatMICE’s ability to leverage spatial patterns dependencies is heavily disrupted when missingness occurs in patterned, more realistic, ways than at random. The KNN algorithm also relies exclusively on spatial information and does not consider temporal proximity. Yet, in contrast to MICE, KNN was among the least affected methods in the shift from random masking to masking by observed patterns. While its MAE still increased by 77.3% under the observed pattern trial, this represented the smallest increase among all methods except for the global mean imputation.

One potential explanation for this discrepancy is that SAITS, SoftImpute, SMILES, and particularly MICE may be more sensitive to structured missingness due to their reliance on a relatively limited set of correlated features during imputation. When these correlated features are frequently missing in the same timesteps, as more often occurs under observed-pattern masking, the models may have access to limited informative inputs which lead to lower performance. Interestingly, this mechanism may not affect KNN as much, since it does not rely more heavily on specific features but instead always uses all the non-missing features in the imputation process.

Examining the results from the masking by observed patterns trial provides some insight into which imputation methods may be best suited for tasks involving ECG data. Among all techniques tested, SAITS emerged as the most consistently accurate. It achieved the lowest overall MAE, outperforming the second-best technique, SMILES, by nearly 9%. Moreover, SAITS-imputed values exhibited the lowest variability in absolute error, indicating not only accuracy but also reliability. Even across different levels of row– or column-wise missingness, SAITS consistently demonstrated superior or at least comparable performance relative to all other imputation methods we evaluated.

However, the benefits of SAITS come with a computational cost. As a deep learning-based model, SAITS requires substantial time and resources to train, particularly when applied to large datasets. In scenarios where computational efficiency is a primary concern, more lightweight methods or simpler alternatives may be appropriate. For instances with lower row-wise missingness (fewer than 40% of missing values), softImpute achieves performance comparable to SAITS. Conversely, when the proportion of missing data per row is higher (i.e., >60% in our analysis), the KNN algorithm generally provides the closest performance to SAITS. Alternatively, when stratified by column-wise missingness, our results suggest using the KNN for columns with less than 40% missing data, and softImpute when missingness exceeds that threshold.

### Clinical Implications

In clinical ECG analysis, missingness is often shaped not by lead disconnection but by whether the signal is interpretable, a distinction with major implications for real-world data pipelines. In our study, missing values reflect instances where our signal processing workflow (a 10-beat median beat averaging approach) failed to produce analyzable output due to excessive noise, arrhythmias, or other signal quality issues. This reflects a common real-world constraint: even when a lead is technically connected, its data may not be usable for feature extraction. Although some methods exist to reconstruct certain leads from others (e.g., usingEinthoven’s law or vectorcardiographic techniques), our study neither employed nor intended to employ such strategies. Instead, we assessed imputation methods in the context of actual feature-level data loss, treating each lead and timepoint independently. This decision aligns with how ECG-derived features are typically handled in research and clinical analytics pipelines, where features are not inferred from each other but instead calculated from directly analyzable signal segments.

However, our reliance on a quality-controlled signal averaging process introduces certain limitations that should inform interpretation of our findings. Because feature extraction depends on forming a valid median beat, some leads or timepoints may exhibit disproportionately high missingness, not due to equipment failure, but due to data quality issues. This introduces a structural bias in the missingness pattern, which may differ from scenarios where missing data arises from electrode detachment or telemetry dropout. Nevertheless, this setup mirrors realistic analytic conditions where waveform noise and arrhythmias are common, and it allows us to benchmark imputation strategies in a way that is both operationally grounded and methodologically transparent. Future research could explore how alternative missingness definitions, such as reconstruction-based imputations or lead correlation modeling, might impact model robustness and downstream analyses. Additionally, future research should also expand the implementation and benchmarking of time-series imputation techniques on raw signals or ECG waveforms to test their clinical usability.

Finally, from a practical standpoint, the feasibility of implementing imputation methods in real-time or near real-time clinical settings depends not only on their accuracy but also on their ability to operate using only past and present information. Several methods we evaluated, such as linear interpolation, the global mean, and KNN, rely on access to the full dataset to perform imputation. Such reliance limits their applicability in real-time prediction or forecasting scenarios, where future values may not be available at the time of decision-making. In contrast, the SAITS model shows promise for real-time deployment due to its temporal awareness and capacity to infer missing values without requiring access to future data. However,SAITS’s advantage must be weighed against themodel’s computational complexity and the infrastructure required for integration into clinical workflows. Future research should explore the imputation capabilities of emerging AI-based models and their potential for real-time clinical decision support.

## 5. Conclusion

In this study, we benchmarked the performance of several imputation methods on pre-processed ECG measurement data using a masking-by-observed-patterns approach (against the conventional random masking), designed to better reflect real-world missingness. Among the methods tested, the recently developed SAITS algorithm consistently achieved the best overall results, with the lowest MAE and lowest variability in imputation error across a variety of missingness scenarios. However, a notable drawback of SAITS is its computational cost; its deep learning architecture can require considerable time to train, particularly on large datasets. In time-sensitive scenarios, simpler methods such as softImpute or KNN may serve as practical alternatives. Our findings suggest that these methods can achieve comparable performance to SAITS under certain conditions, such as when missingness is relatively low by row (favoring softImpute) or column (favoring KNN).

Beyond comparing imputation performance, this work highlights a critical methodological concern, which is the way in which missingness is introduced during benchmarking significantly influences error estimates. The current standard practice for benchmarking involves applying a *random mask* to observed values and then computing error metrics using the known ground truth. However, when we compared this approach to a more realistic alternative (*masking based on observed missingness patterns),* it became clear that random masking may substantially underestimate the imputation error. Specifically, random masking does not fully capture the real-world challenges that imputation algorithm must overcome, such as long consecutive missing values (gaps) within individual features or simultaneous absence of correlated variables at a given time step. These characteristics, which are often present in naturally missing ECG data, can severely disrupt both temporal and spatial imputation strategies. As such, relying solely on random masking for performance evaluation may yield overly optimistic assessments of an algorithm’s effectiveness in practice.

Nonetheless, in many circumstances, using observed-pattern masking for benchmarking may not always be feasible. This masking approach requires access to source samples with very low natural missingness, which may not be available in many real-world datasets. In such cases, practitioners or researchers may be forced to rely on random masking, despite its limitations. This underscores the need for further research into benchmarking strategies that better simulate realistic missingness while remaining broadly applicable. Future work should also focus on developing an improved masking approach that preserves critical structural characteristics of naturally missing data without requiring access to rare complete records.

## Funding Acknowledgment

This work was funded by the University of Rochester Office of the Vice President for Research, the School of Medicine and Dentistry and Arts, Sciences, & Engineering via the Center for Integrated Research Computing (PI: Suba). The COMPARE Study was supported by grant R21NR011202 (PI: Pelter) provided by the National Institutes of Health.

## Data Availability

Aggregated data (e.g. summary tables, processed feature-level values) of the ECG that support the findings in this manuscript can be made available upon reasonable request. Codes used in our analysis are available at GitHub: https://github.com/suksuba/ecg-imputation.

