## Supplement Figures for "Time-series ECG Imputation Using a Pattern-Based Masking Framework"

*[1] School of Nursing, University of Rochester Medical Center, Rochester, NY; [2] Goergen Institute for Data Science and Artificial Intelligence, University of Rochester, Rochester, NY; [3] Clinical Cardiovascular Research Center, University of Rochester Medical Center, Rochester, NY; [4] Department of Physiological Nursing, School of Nursing, University of California San Francisco, San Francisco, CA*

### *Supplement Figures Captions*

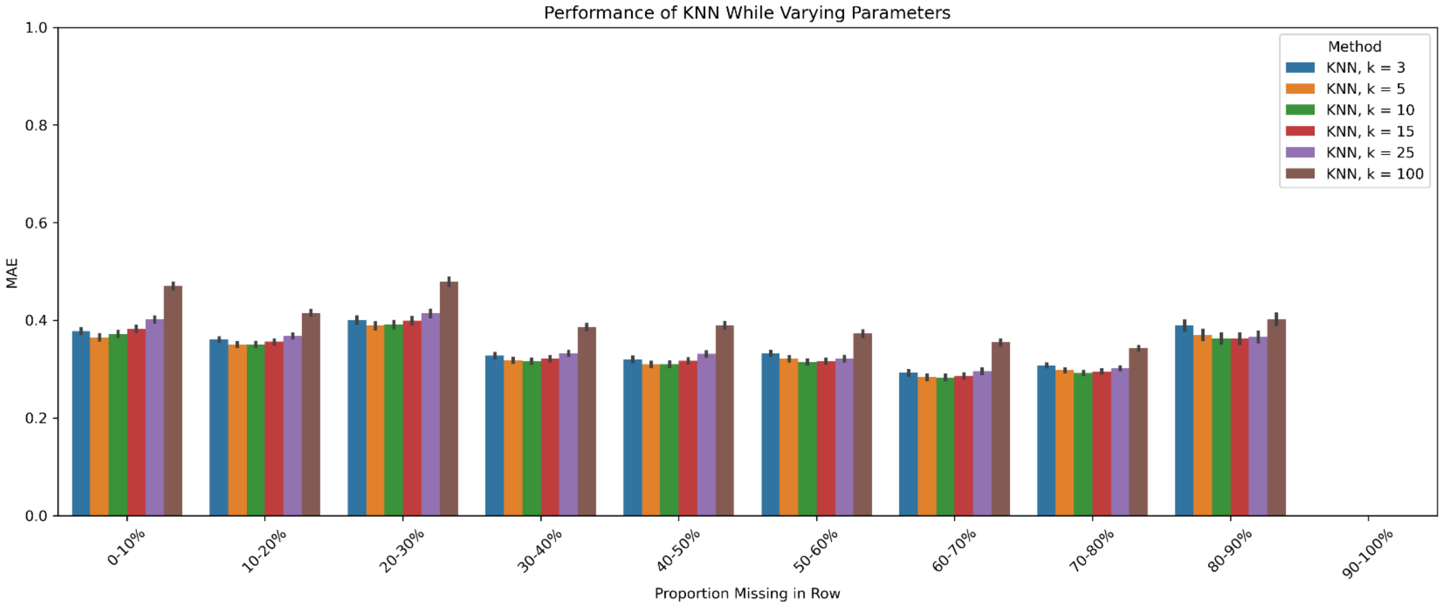


**Suppl. Fig 1.** The performance (mean absolute error) of the K-Nearest Neighbor (KNN) method using different values of the *k* parameter at varying levels of data missingness by row.

¶ These authors contributed equally to this work.

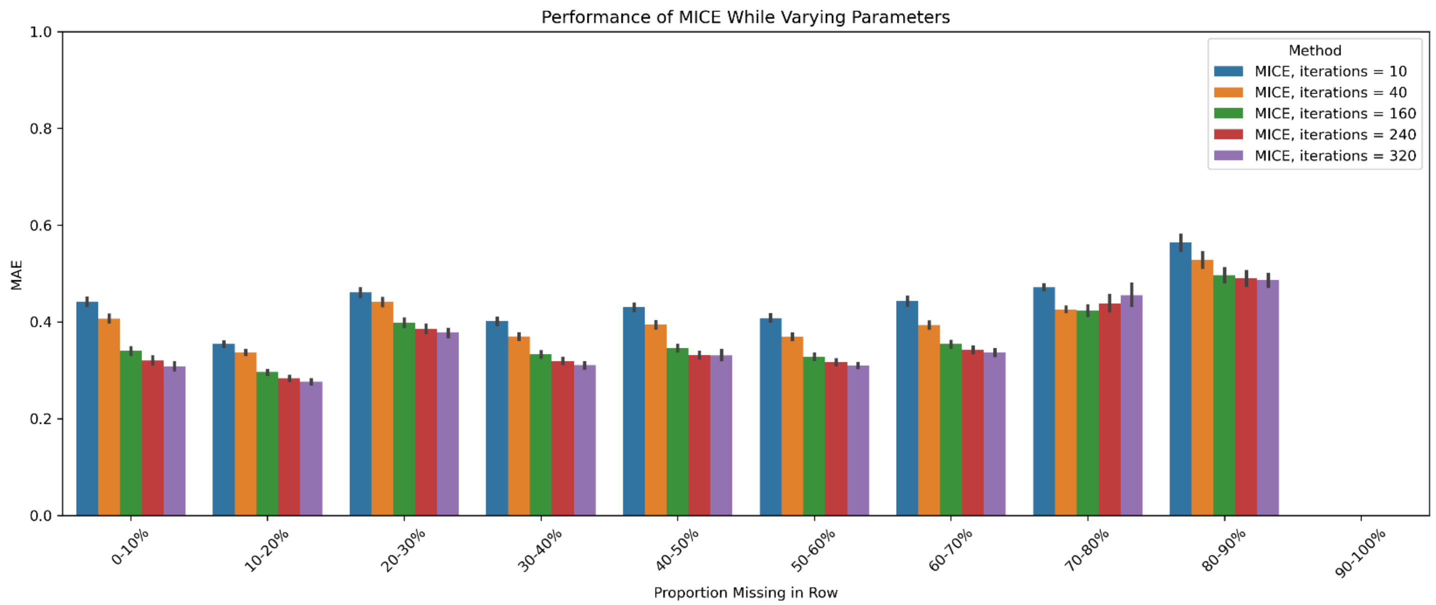


**Suppl. Fig 2.** The performance (mean absolute error) of the Multivariate Imputation via Chained Equations (MICE) method for different values of the max iterations parameter at varying levels of data missingness by row.

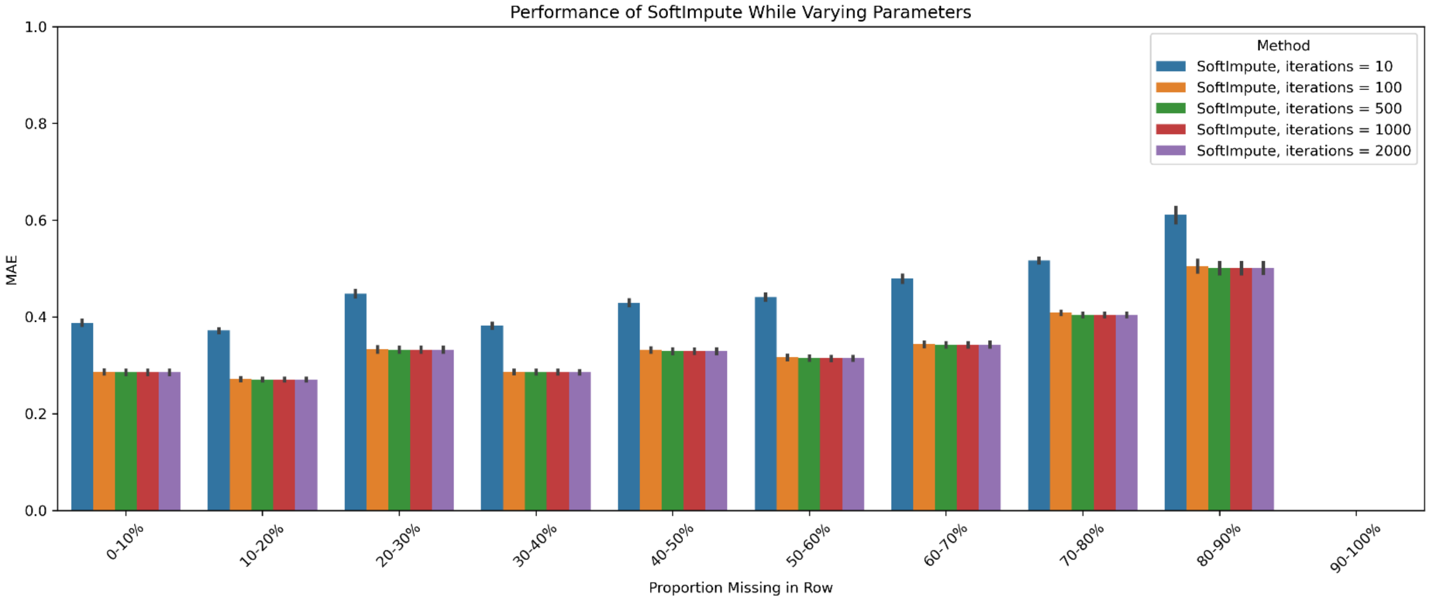


**Suppl. Fig 3.** The performance (mean absolute error) of the SoftImpute component of the SMILES (xgbooSt MIssing vaLues In timE Series) method for different values of the max iterations parameter at varying levels of data missingness by row.
